# Observational study on the clinical epidemiology of infectious acute encephalitis syndrome including Nipah virus disease, Bangladesh: BASE cohort study protocol

**DOI:** 10.1101/2025.04.30.25326714

**Authors:** Md Zakiul Hassan, Amanda Rojek, Dewan Imtiaz Rahman, Sharmin Sultana, Mahbubur Rahman, Mohammad Khaja Mafij Uddin, Mohammad Enayet Hossain, Md Wasik Rahman, Laura Merson, Esteban Garcia, Jake Dunning, Josephine Bourner, Shadman Sakib Choudhury, Kamal Ibn Amin Chowdhury, Khalequ Zaman, Md. Sharful Islam Khan, Mohammad Hasan Tarik, Shahida Yeasmin, Faruk Ahammad, Abu Faisal Md Pervez, Md Mahfuzer Rahman, Abu Rayhan Md Suja-Ud-Doula, John D. Klena, Christina F. Spiropoulou, Trevor Shoemaker, Sayera Banu, Joel M Montgomery, Mohammed Ziaur Rahman, Tahmina Shirin, Syed M Satter, Peter Horby, Piero Olliaro

## Abstract

**Background:** Nipah virus (NiV) is a bat-transmitted paramyxovirus causing recurrent, high-mortality outbreaks in South and South-East Asia. As a WHO priority pathogen, efforts are underway to develop therapies like monoclonal antibodies and small-molecule antivirals, which require evaluation in clinical trials. However, trial design is challenging due to limited understanding of NiV’s clinical characteristics. Given the rarity of NiV infections, strategies targeting improved outcomes for the broader acute encephalitis syndrome (AES) patient population, including those with NiV, are essential for advancing therapeutic research. To address these gaps, we designed the Bangladesh Acute Encephalitis Syndrome (BASE) cohort study to characterise the patient population, clinical features, treatment practices, common aetiologies and outcomes in patients presenting with AES, including NiV infection, to inform the design of clinical trials for NiV and AES more broadly.

**Methods:** This prospective cohort study will be conducted in Bangladesh, a NiV endemic country with annual outbreaks. In collaboration with the ongoing NiV surveillance programme in Bangladesh, we aim to enrol up to 2,000 patients of all ages presenting with AES at three tertiary care hospitals within the Nipah belt. Patients who provide informed consent to participate will be monitored throughout their hospital stay until 90 days post-discharge. Data will be systematically collected through interviews and medical record reviews at several time points: on the day of enrolment, day 3, day 7, the day of critical care admission (if applicable), discharge day, and 90 days post-discharge. Additionally, a portion of the cerebrospinal fluid (CSF) collected under the concurrent NiV surveillance protocol will be tested for an array of viral and bacterial pathogens responsible for encephalitis at the International Centre for Diarrhoeal Disease Research Bangladesh (icddr,b) laboratory.

**Discussion:** By characterizing the AES patient population, this study will generate essential evidence on key clinical parameters, which will be pivotal in optimizing the design of clinical trials for potential interventions aimed at improving outcomes in patients with AES, including those with NiV disease.

**Clinical trial number:** not applicable (this is an observational study)

## 1 Background

Nipah virus (NiV) is a highly lethal zoonotic paramyxovirus transmitted through contact with infected bats, pigs, humans, or consumption of contaminated raw date palm sap (1, 2). It causes severe neurological and respiratory illness with a high mortality rate (3, 4). Since its discovery in 1998/1999 in Malaysia and Singapore, outbreaks have occurred in Bangladesh, India, and the Philippines, with concerns about its spread to regions where *Pteropus* bats, the virus’ natural reservoir, are found (5, 6). Mortality has remained consistently high across outbreaks, and there are no approved treatments (7, 8). Current management relies on supportive care, but delays to diagnosis, and the lack of standardized protocols and limited access to intensive care in affected regions pose major challenges (9). Strengthening clinical management and optimizing interventions through data-driven approaches are critical to prepare for larger future epidemics.

NiV disease (NiVD) typically manifests as acute encephalitis syndrome (AES), with or without pulmonary disease (10). AES carries substantial morbidity and mortality, particularly impacting individuals living in low- and middle-income countries, including those also affected by NiVD (11). Reports from India, a Nipah endemic country, showed an all-cause encephalitis mortality ranging from 14% to 36% (12, 13). Reviews on AES outcomes revealed that nearly half of surviving children experienced neurodevelopmental sequelae, including developmental delay and motor impairment (14). In adults, 26% to 62% faced significant long-term consequences, including epilepsy, memory issues, inappropriate behaviour, social skill deficits, fatigue, personality changes, cognitive problems, and difficulties in daily living skills (15).

Diagnosing AES remains a significant challenge, with up to 85% of cases globally undiagnosed, even in high-income settings with advanced diagnostic capabilities, due to the need for extensive sampling and complex analyses (16-20). Viruses such as herpesviruses, arboviruses, enteroviruses, and adenoviruses are primary etiological agents, while bacterial pathogens including *Rickettsia* species and *Mycobacterium tuberculosis* (MTB) also contribute in certain regions; fungal and parasitic infections are less common (21-23). In countries like Bangladesh and India, where NiVD is endemic, AES diagnosis is further complicated. NiVD, a relatively rare cause of AES (approximately 3% of AES cases in Bangladesh), often presents with nonspecific symptoms overlapping with those of other common AES pathogens (22-24). Clinically indistinct symptoms combined with limited diagnostic access, especially in rural areas, increases diagnostic uncertainty and highlights limitations within current AES diagnostic protocols.

To gain a comprehensive understanding of the clinical epidemiology of NiVD, including neurological and pulmonary involvement, a prospective observational study is needed. Clinical characterisation of NiVD disease including the broader group of patients with AES should form the foundation for developing standardized clinical trial methodologies, including enrolment criteria, outcome measures, and subgroup analyses. The clinical epidemiology of AES can also facilitate the identification of high-risk patients, optimizing the allocation of diagnostic resources and enhancing overall disease management strategies. This involves improving contact tracing and implementing infection prevention and control (IPC) protocols within hospitals, especially in settings with limited resources.

Enhanced clinical epidemiological studies should also document current clinical care practices to identify gaps, inform the development of context-specific standard of care guidelines, and to define the baseline standard of care for clinical trials. During the 1998-99 Malaysian outbreak, 10 out of 11 NiV patients cared for in a Singapore hospital survived (25), suggesting that improving elements of supportive care improves patient outcomes. Currently, there are several potential NiV-specific therapeutics in different phases of development, these include monoclonal antibodies and small molecules antivirals (26). By characterizing the range of causes of AES, we will generate evidence on key clinical parameters necessary to inform the design of clinical trials for potential interventions aimed at improving clinical outcomes in the broader group of patients with AES, including those with NiVD.

## 2 Aim and objectives

The aim of this study is to describe the patient population, clinical presentation, natural history, common infectious aetiologies, treatment practices, and clinical outcomes of patients presenting with AES (including NiVD) to inform the design of future clinical treatment trials.

The primary and secondary objectives of the study with relevant outcome measures are listed in Table 1.

**Table 1.**
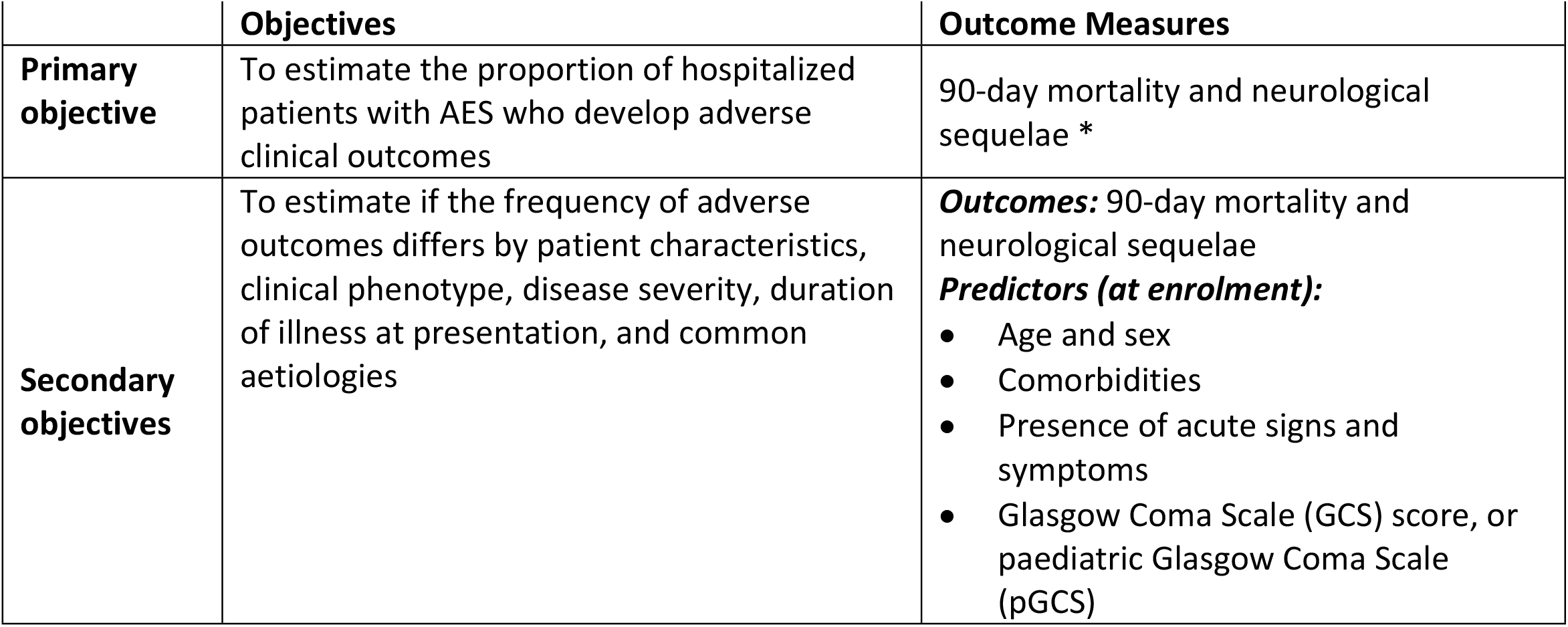
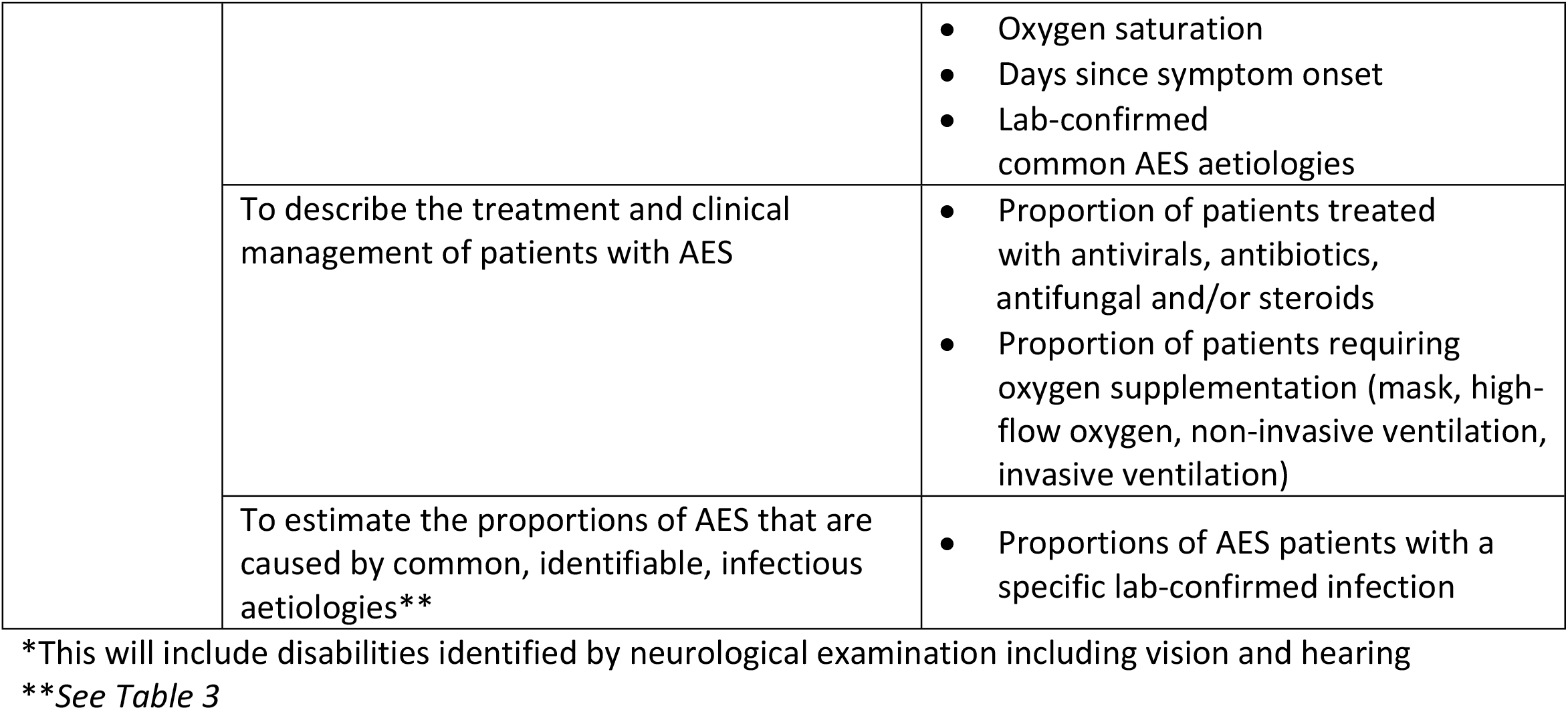
Study objectives and relevant outcome measure.

|  | **Objectives** | **Outcome Measures** |
| --- | --- | --- |
| **Primary objective** | To estimate the proportion of hospitalized patients with AES who develop adverse clinical outcomes | 90-day mortality and neurological sequelae * |
| **Secondary objectives** | To estimate if the frequency of adverse outcomes differs by patient characteristics, clinical phenotype, disease severity, duration of illness at presentation, and common aetiologies | **Outcomes:** 90-day mortality and neurological sequelae<br>**Predictors (at enrolment):**<br>• Age and sex<br>• Comorbidities<br>• Presence of acute signs and symptoms<br>• Glasgow Coma Scale (GCS) score, or paediatric Glasgow Coma Scale (pGCS) |
|  |  | • Oxygen saturation<br>• Days since symptom onset<br>• Lab-confirmed common AES aetiologies |
|  | To describe the treatment and clinical management of patients with AES | • Proportion of patients treated with antivirals, antibiotics, antifungal and/or steroids<br>• Proportion of patients requiring oxygen supplementation (mask, high-flow oxygen, non-invasive ventilation, invasive ventilation) |
|  | To estimate the proportions of AES that are caused by common, identifiable, infectious aetiologies** | • Proportions of AES patients with a specific lab-confirmed infection |

\*This will include disabilities identified by neurological examination including vision and hearing
\*\*See Table 3

## 3 Study design

The Bangladesh Acute Encephalitis Syndrome (BASE) study is a hospital-based prospective observational cohort study. Patients who meet the eligibility criteria will be invited to provide informed consent to participate. Enrolled patients will have demographic and clinical data collected during hospital admission and will be followed up at several specific time points up until 90 days after enrolment.

The study will be integrated with clinical care and with an ongoing surveillance programme titled ‘Nipah Virus Transmission in Bangladesh’ jointly implemented by the International Centre for Diarrhoeal Disease Research Bangladesh (icddr,b) and the Institute of Epidemiology, Disease Control, and Research (IEDCR), Bangladesh with technical support from the United States Centers for Disease Control and Prevention (CDC) (27). Clinical staff, surveillance staff, and BASE study staff will collaborate to align procedures to minimise any burden to participants.

### 3.1 Routine clinical assessment and investigation of encephalitis patients

Patients admitted to hospital with AES will have a clinical examination, medical history, laboratory investigations and imaging based on the suspected diagnosis and local availability of tests. Routine investigations may include a complete blood count (CBC), liver function tests (LFTs), renal function test (RFT), electrolytes, blood glucose, and cerebrospinal fluid (CSF) examination. Imaging such as Computed Tomography (CT) and Magnetic Resonance Imaging (MRI) of the brain may also performed to aid in diagnosis and treatment planning. The specific tests conducted for each patient depends on factors like the suspected diagnosis, patient preferences, and the patient’s willingness or ability to cover the costs. For similar reasons, not all patients undertake routine cerebrospinal fluid (CSF) testing, even though it is recommended for AES patients by local guidelines by the IEDCR (9).

When available, blood and CSF specimens are tested by the hospital laboratory. Imaging studies are conducted either at the hospital’s facility or, if unavailable, at a nearby private facility. Clinical data, investigation reports, and treatment details are recorded in a paper format as public hospitals currently lack an electronic medical record system. Clinical care data will be recorded in the BASE study electronic case report form as per the study schedule.

### 3.2 Nipah surveillance programme activities

We will leverage the ongoing NiVD surveillance programme for participant recruitment (27). The national NiVD surveillance programme has been active across Bangladesh since 2007. The three hospitals reporting the most NiVD cases over the past 15 years will implement the BASE study alongside the surveillance programme. All AES cases identified through the surveillance programme at these three sites will be informed of the BASE study and invited to participate. In addition, any confirmed NiVD cases identified through the surveillance programme during the study period but occurring outside of the three BASE study hospitals will also be invited to participate in BASE study.

Daily screening of all patients admitted to the emergency and inpatient departments will be done by the surveillance team to identify patients. When a patient is identified through surveillance screening (AES at the three main hospitals or NiV infection at any other hospital), a surveillance programme team member will invite the patient or their relative/guardian to provide informed consent. Data will be captured on those who agree, and a throat swab and blood sample will be collected. If a sample of CSF will be taken for clinical care, consent to test an aliquot of this sample will be requested as a part of the surveillance programme.

### 3.3 Study sites

The main study sites are three tertiary care public hospitals Who are engaged with the ongoing NiV surveillance programme located in the NiVD belt (Figure 1):

**Figure 1:**
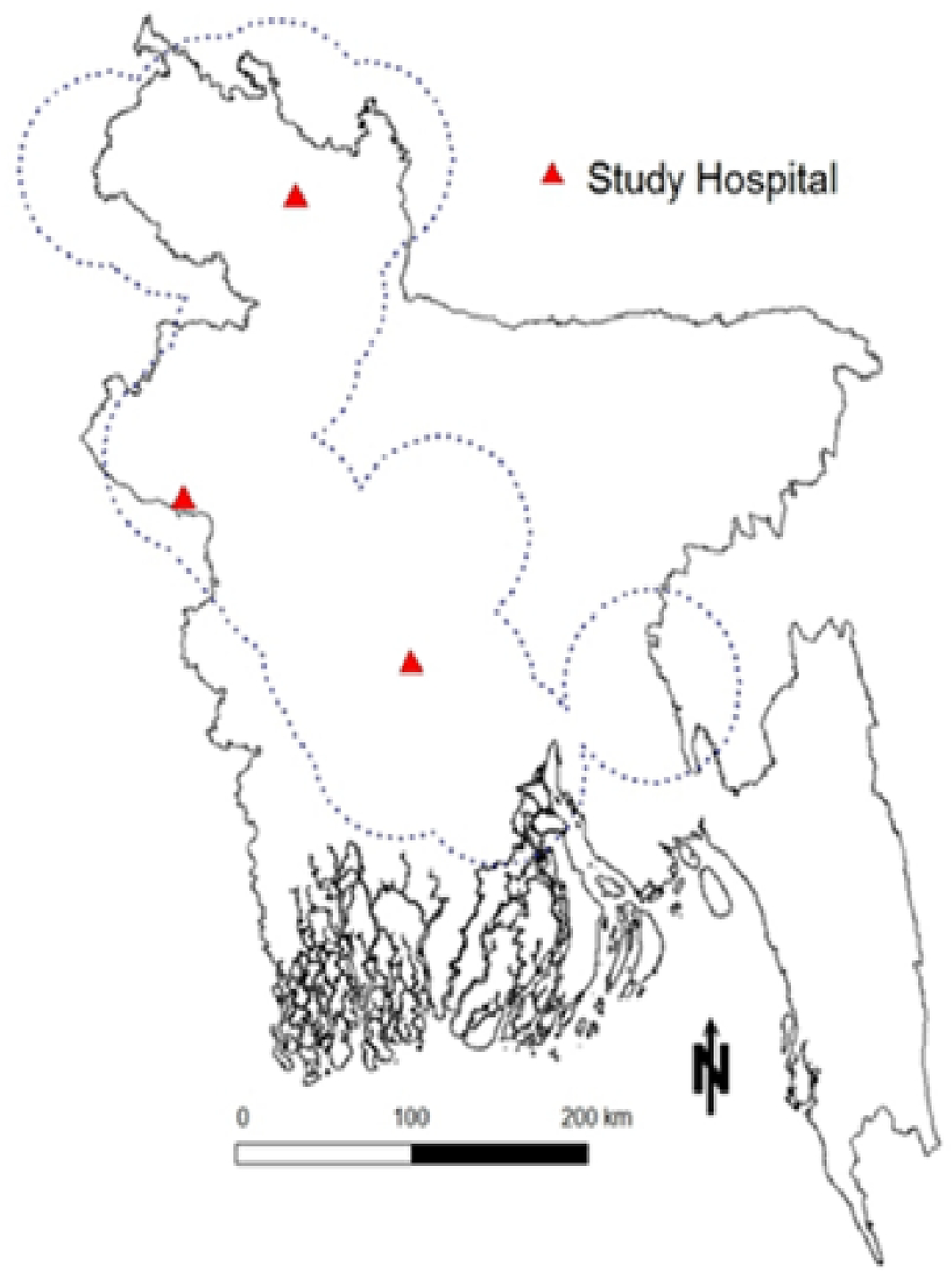
Location of main BASE study sites, the dotted line indicates the Nipah belt within Bangladesh.

1. Rajshahi Medical College Hospital, Rajshahi
2. Rangpur Medical College Hospital, Rangpur
3. Bangabandhu Sheikh Mujib Medical College Hospital, Faridpur

The hospitals have been chosen primarily based on their location within the Nipah belt and their record of reporting the most NiVD cases. They also ensure diverse geographical representation, have high bed capacity, manage high patient volumes, and are equipped with facilities for routine laboratory tests such as CSF examination. Additionally, patients with confirmed NiVD diagnosis at any hospital in Bangladesh will be alerted to the surveillance programme and, subsequently, BASE study staff for potential recruitment. Study staff based at the main sites will travel to sites across the country when alerted of a patient with confirmed NiV infection. Over the past 20 years, an average of 14 cases of NiVD have been reported per year in Bangladesh (10). Though unlikely, additional NiVD cases will be enrolled if a larger outbreak occurs.

### 3.4 Study population

AES patients of any age and sex admitted to study hospitals and meeting the inclusion criteria are eligible to enrol. In addition, all laboratory confirmed NiVD cases identified by NIVD surveillance programme during the study period, regardless of location of clinical care, are eligible to enrol.

### 3.5 Inclusion Criteria

We will use the enrolment criteria used by the NiVD surveillance programme. Patients of any age and sex admitted with ***acute encephalitis*** defined as:

1. Reported or measured fever (axillary temperature >38.5°C) **AND**
2. Evidence of acute brain pathology (e.g., altered mental status, new onset seizures, or new neurological deficit either diffuse or localized to the brain).
  - Participant is enrolled in the icddr,b-IEDCR NiV surveillance programme
  - Participant or their legal representative/guardian is willing and able to provide informed consent for participation in the study

### 3.6 Exclusion Criteria

The participant may not enter the study if they have a clear alternative non-infectious disease diagnosis (either clinical or laboratory/imaging confirmed diagnosis) that explains the acute presentation.

### 3.7 Screening and eligibility assessment

For potential participants, enrolment in the NiVD surveillance programme will be an inclusion criterion for the BASE study due to reliance on the programme’s NiV testing which is the primary pathogen of interest. Surveillance programme staff will inform the BASE study team of all patients who consent to enrol in the NiVD surveillance program in the three study hospitals and of any confirmed NiVD patients located at any hospital. BASE study staff will verify the eligibility of patients against the study inclusion and exclusion criteria and initiate a discussion with the patient or their parent/guardian/representative regarding the BASE study. An anonymised screening log tracking the number of patients who meet or do not meet the enrolment criteria, and who consent or do not consent to the BASE study will be maintained. Anonymised information regarding the number of patients screened and enrolled to the surveillance programme will be requested from the surveillance programme staff to inform understanding of complete AES patient numbers at each site.

### 3.8 Enrolment

Participants will be enrolled in the study after written informed consent has been obtained and their enrolment has been registered on the electronic case record form (eCRF). An enrolment log including patient names and contact details will be completed and securely stored at the study site.

### 3.9 Recruitment status

Participant recruitment began in March 2023 and is ongoing. Recruitment will continue until the target of 2,000 participants is reached. As of April 2025, over half of the targeted participants have been enrolled across the three study sites. Recruitment is expected to conclude by the end of 2025. Data collection, including follow-up assessments at 90 days post-discharge, will continue through the first quarter of 2026. Final study results are anticipated to be available by the second quarter of 2026.

### 3.10 Data collection

Clinical and laboratory information will be documented on enrolment, day 3, day 7, upon admission to critical care (if applicable), and at hospital discharge. Clinical information collected between admission and enrolment will also be collected. Clinical and laboratory information will be gathered from the hospital records, patient examination/discussion, and surveillance programme records. Study staff will record all laboratory test values and imaging results done as part of the routine care such as Complete Blood Count (CBC), liver function tests (LFTs), renal function tests (RFTs), electrolytes, CSF examination and imaging such as Chest x-ray, CT scan, and MRI (see Table 2 below).

**Table 2:** Schedule of study procedures.

|  | Day 1 | Day 3 | Day 7 | ICU Admission | All available routine investigation | Discharge | Day 90 |
| --- | --- | --- | --- | --- | --- | --- | --- |
|  | Enrollment | In hospital (or by telephone if discharge before day 7) |  |  |  |  | Outpatients follow-up |
| Eligibility screen | × |  |  |  |  |  |  |
| Informed consent | × |  |  |  |  |  |  |
| Data collection - clinical features |  |  |  |  | × |  |  |
| Demographic | × |  |  |  |  |  |  |
| Exposure | × |  |  |  |  |  |  |
| Baseline clinical information | × |  |  |  |  |  |  |
| Physical examination | × | × | × | × |  |  |  |
| Clinical observation | × | × | × | × |  | × |  |
| Treatment and interventions | × | × | × | × |  | × |  |
| Complications | × | × | × | × |  | × |  |
| Outcome |  |  |  |  |  | × | × |
| Neurological assessment |  |  |  |  |  | × | × |
| Laboratory testing |  |  |  |  |  |  |  |
| Testing of available routine or surveillance CSF aliquot for target pathogens | × (1) |  |  |  |  |  |  |
| Data collection - available routine laboratory record |  |  |  |  |  |  |  |
| CBC | × |  |  |  | × |  |  |
| Blood glucose |  |  |  |  | × |  |  |
| CRP | × |  |  |  | × |  |  |
| Liver function tests | × |  |  |  | × |  |  |
| Renal function | × |  |  |  | × |  |  |
| Electrolytes | × |  |  |  | × |  |  |
| CSF | × |  |  |  | × |  |  |
| Data collection - available routine imaging records |  |  |  |  |  |  |  |
| Chest X-Ray | × |  |  |  | × |  |  |
| MRI of Brain | × |  |  |  | × |  |  |
| CT Brain | × |  |  |  | × |  |  |
| Data collection - available routine or surveillance programme microbiological tests |  |  |  |  |  |  |  |
| Nipah virus | × |  |  |  | × |  |  |

(1) If CSF sample taken for clinical care or surveillance programme

If CSF has been taken as a part of clinical care, the hospital laboratory will provide an aliquot to be tested for infectious pathogens by the study laboratory at icddr,b.

If permission from the patient is given, patients who are discharged before day 7 may have a phone call from the study team to request that the clinical information from the day 3 and/or 7 data collection dates be included. Patients will be invited to attend a follow-up visit on Day 90. These visits will take place at one of the three study hospitals, or, if more convenient for the patient, at a nearby hospital where the patient is located or as a home visit if the patient is unable to travel. The visit will include a clinical assessment and a neurological assessment, including vision and hearing testing, that will take approximately one hour. Patients who are unable or unwilling to attend the follow-up visit in one of the hospitals or at home will be invited to have a telephone consultation to collect follow-up data.

Patients who report to be pregnant at any point during the study will be asked if the study team may contact them to know the outcome of their pregnancy.

### 3.11 Sample processing, transportation and laboratory testing

Data will be collected on the results of samples and laboratory analysis that are part of routine clinical care or the NiVD surveillance programme. If CSF specimens are collected for routine care or NiVD surveillance, an aliquot will be stored for infectious pathogen testing at icddr,b (Table 3). Aliquots made for this study will depend on availability of volume. Clinical and surveillance laboratory processing will take precedence in the absence of sufficient volume for all analyses. Samples will be handled and shipped by the hospital laboratory according to routine procedures. Shipments will be made to the icddr,b laboratory in Dhaka where processing and testing will follow established protocols.

**Table 3:**
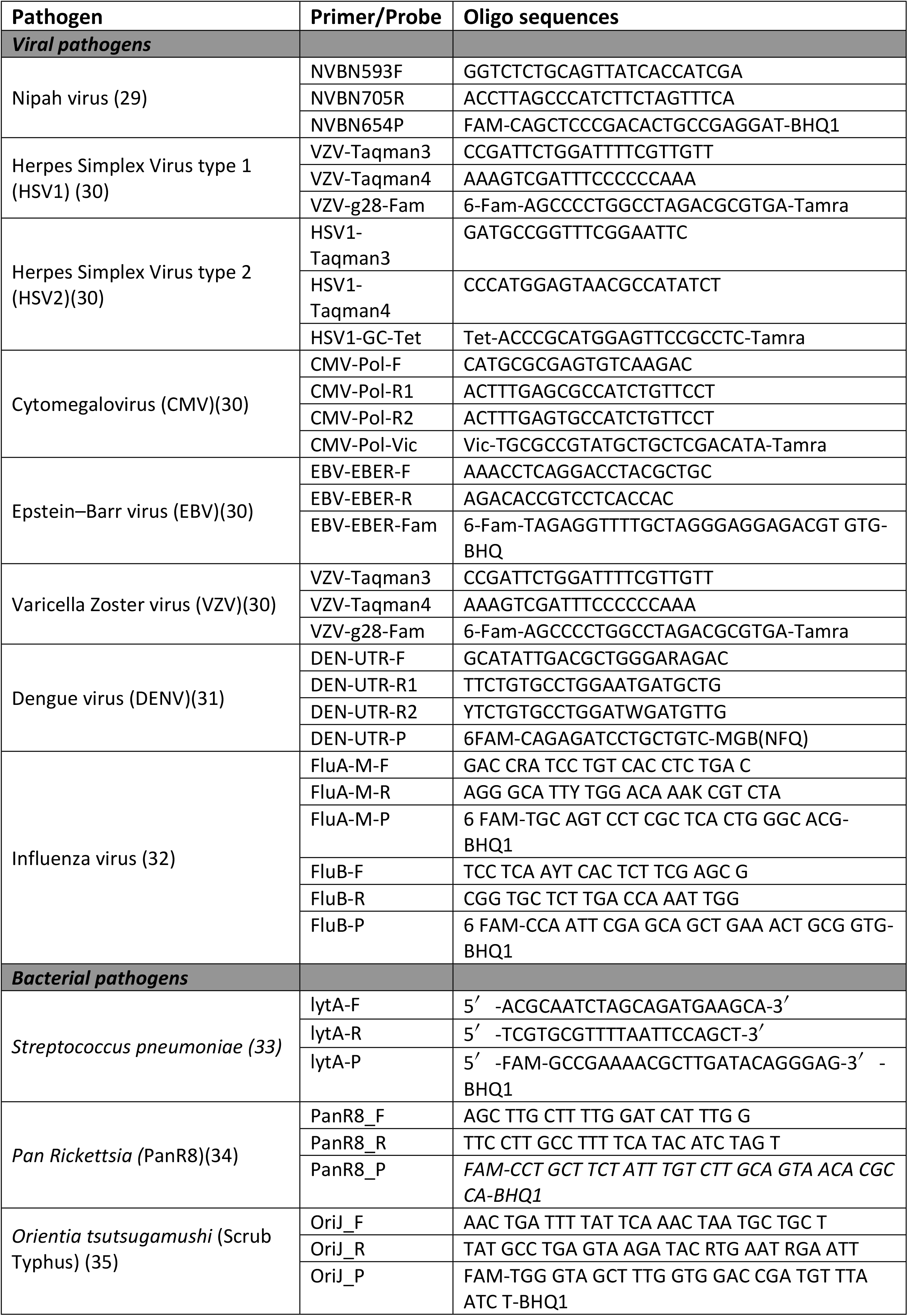
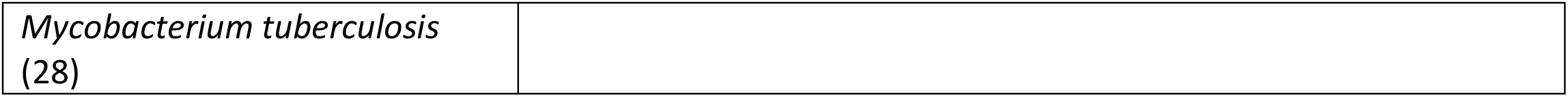
List of target pathogens tested by real time RT-PCR.

In brief, an aliquot of CSF will be transferred into lysis buffer (NucliSENS easyMag, bioMerieux Inc., Rodolphe St., Durham, NC, USA)) to inactivate potential high-risk pathogens while stably preserving nucleic acids, ensuring the specimens are safe for handling in a BSL-2 environment. Total nucleic acids from the CSF will be extracted using a InviMag Virus DNA/RNA Mini Kit (INVITEK Molecular, Berlin-Buch GmbH, Germany) on Kingfisher Flex 96 (Thermo Fisher Scientific Inc., Waltham, MA, USA) automated nucleic acid extraction system according to the manufacturer’s instruction. The eluted NA will be of high quality and suitable for downstream molecular analyses.

Real-time reverse transcription polymerase chain reaction (rRT-PCR) assays will be conducted using commercially recommended kits specific for both DNA and RNA targets (Table 3). The mastermix will be prepared following the manufacturer’s guidelines, and the template will be added accordingly. The rRT-PCR assays will be performed on a BioRad CFX-Opus PCR system (BioRad, Hercules, California, USA) using the recommended thermal profile provided by the assay developer. Positive and negative controls will be included in each rRT-PCR experiment to ensure assay accuracy. Amplification curves will be visually checked, and tests with poor-quality curves will either be repeated or considered negative. A cycle threshold (CT) value of <35 will be classified as positive, while values ≥35 will be regarded as negative.

For the rapid diagnosis of Mycobacterium tuberculosis, we will use the GeneXpert MTB/RIF Ultra test (Xpert Ultra), an advanced, cartridge-based nucleic acid amplification test (28). The Xpert Ultra, recommended by the WHO, is specifically designed to detect both Mycobacterium tuberculosis and rifampicin resistance. Utilizing advanced melting curve analysis, this next-generation assay targets a key region of the rpoB gene, including its critical 81-base pair “core region,” as well as sequences in the multi-copy IS1081 and IS6110 insertion elements. This enhanced sensitivity and specificity make the Xpert Ultra an optimal tool for prompt and reliable tuberculosis diagnosis and drug resistance testing.

In cases where multiple pathogens are identified in a sample, an expert panel consisting of microbiologists, epidemiologists, and clinicians will review the patient’s clinical, laboratory, and exposure records to determine the most likely causative pathogen. Test results will be regularly reported to the respective hospitals to aid in patient management.

### 3.12 Sample size determination

This is a descriptive observational study aimed at characterizing AES, including NiVD. Our aim is to collect detailed clinical data, given our limited understanding of the clinical characteristics of AES. The primary outcome of interest (to inform clinical trial design) is the proportion of AES patients with an adverse clinical outcome (mortality or neurological sequelae). To ensure the study captures sufficient number of outcome events to provide informative and representative data, we estimated a sample size based on the two primary outcomes of interest (mortality and neurological sequelae) to guide required enrolment.

Data from ongoing NiVD surveillance in Bangladesh during 2022-2023 showed an in-hospital mortality rate of around 15% (756/4952) of enrolled AES participants (icddr,b-IEDCR NiVD surveillance team, personal communication). Studies from neighbouring India also reported an all-cause 14%-36% mortality rate among hospitalized encephalitis patients irrespective of aetiologies (12, 13).

Assuming an expected mortality outcome of 15%, the estimated required sample size is 1532 for the margin of error or absolute precision of ±2% in estimating the case fatality ratio with 95% confidence and considering the potential loss/attrition of 20%. With this sample size, the anticipated 95% CI is (13%, 17%). This sample size is calculated using the formula for estimating a proportion as previously described (36). The reported prevalence of long-term neurological sequelae varied between 32%-53% for children and 26%-62% for adults depending on the duration of follow-up (14) (11). Therefore, for the long-term neurological sequelae outcome, we assume a conservative prevalence of 20% for long-term neurological sequelae outcome of enrolled participants during our 90-day follow-up. With 20% prevalence the estimated required sample size is 1922 for the margin of error or absolute precision of ±2% in estimating the prevalence of long-term neurological sequelae with 95% confidence and considering the potential loss/attrition of 20%. With this sample size, the anticipated 95% CI is (18%, 22%).

To capture the required number of both outcome events, we plan to enrol 2000 AES patients in our study. We plan to conduct the study in three purposively selected tertiary care hospitals that have ongoing NiVD surveillance and are situated within the Nipah belt. The enrolment of patients will vary across sites due to differences in facility size and catchment area.

### 3.13 Statistical analysis

We will perform descriptive analysis of patient characteristics, acute signs and symptoms, laboratory parameters, disease severity, common aetiologies, and clinical management, including medications and clinical outcomes of patients. Continuous data will be presented as a mean with Standard Deviation where data are normally distributed and as a median with the 25th and 75th centiles for non-parametric data along with a 95% confidence interval for both the mean and the median Categorical data will be summarised as frequencies and percentages.

We will also compare frequency of adverse outcomes (mortality and long-term neurological complications categorized by severity: mild, moderate, or severe) by patient characteristics, exposures (date palm sap consumption, contact with infected humans, bats, or pigs), acute signs and symptoms, disease severity, duration of illness at presentation, and common aetiologies. For univariate comparisons, Welch’s t-test for two groups or ANOVA for more than two groups will be used when dealing with normally distributed continuous data. Non-parametric continuous data will be compared using the Mann-Whitney U test for two groups or the Kruskal-Walli’s test for three or more groups. In cases of categorical data, differences will be assessed using the χ^2^ test, or Fisher’s exact test where applicable.

To measure the association between patient characteristics, acute signs and symptoms, disease severity, duration of illness at presentation, and common aetiologies, and adverse outcomes defined as 90-day mortality and neurological sequelae, we will calculate hazard ratios using multivariable Cox regression after adjusting for potential confounders.

A missing at random assumption (MAR) will be made for missing data however this will be investigated to determine whether reasons for missing data can be obtained and to determine if such an assumption is realistic. Patterns of missing data across sites, and patient characteristics will be reported. In this study, we will address the issue of missing data using Multiple Imputation, which is particularly adapted to data that are Missing Completely at Random (MCAR) or Missing at Random (MAR). This method involves creating multiple complete datasets by imputing missing values, analysing each dataset separately, and then pooling the results to account for the uncertainty caused by the missing data. This approach aims to preserve statistical power and reduce bias in the analyses. Additionally, sensitivity analyses will be conducted to evaluate the robustness of our results under different assumptions about the missing data. This strategy should ensure that the conclusions reached are reliable and reflective of the true effects, despite the presence of missing data.

### 3.14 Protection of human subjects and ethical approval

The BASE study will be conducted in strict adherence to ethical standards, ensuring the confidentiality of all participants in accordance with applicable Good Clinical Practice (GCP) guidelines, national regulations, and the Declaration of Helsinki. Informed consent will be obtained from all participants or their legal representatives prior to enrolment. The consent process will clearly explain the study’s purpose, procedures, potential risks and benefits, and the right to withdraw at any time without affecting future care. The consent form will be provided in Bengali, and participants will have ample opportunity to ask questions before and after deciding on participation. For those unable to read or write, the consent process will include an impartial witness to confirm comprehension and voluntary participation.

To protect participant confidentiality, data will be pseudonymized, with each patient assigned a unique patient identification number, which will be consistently used throughout the study. This number will be assigned and managed securely by the centre coordinator, and personal data will only be released in accordance with local regulations and ethics committee approvals. The study received ethical approval from the Oxford Tropical Research Ethics Committee (OxTREC Ref: 576-23) and the Institutional Review Board (Research Review and Ethics Review Committee) of icddr,b (icddr,b Protocol Number: 24016). Any future amendments to the study protocol will also be submitted for review and approval to these committees.

## 4 Discussion

The BASE study is designed to address critical gaps in the clinical understanding of AES, including NiVD, and to provide essential data that will inform future clinical trials aimed at evaluating potential treatments for NiVD. AES, a complex syndrome with multiple infectious causes, poses significant diagnostic and therapeutic challenges, particularly in South and Southeast Asia, where NiV is endemic. Given the small numbers of NiVD cases, conducting large-scale, disease-specific trials is not feasible. Therefore, the BASE study adopts a syndromic approach, integrating NiV within the broader AES context to allow the evaluation of potential interventions in a more practical and inclusive framework. This design not only facilitates the collection of critical data on NiV but also enhances the understanding of AES management overall.

The study will generate crucial clinical insights by systematically collecting detailed data on patient demographics, clinical presentations, disease severity, treatments, and outcomes. This includes information on age, sex, comorbidities, oxygen saturation levels, GCS scores, patterns of organ involvement, and duration of illness. These data are essential for informing future clinical trials, as they will help determine how these factors influence patient outcomes. For instance, patient demographics and comorbidities can guide stratification strategies to ensure balanced treatment groups, while detailed clinical presentations and disease progression will help define disease phenotypes and identify predictors of adverse outcomes (**Table 4**). Additionally, understanding the timing of symptom onset and treatment response will aid in determining optimal therapeutic windows, ensuring that interventions are applied at the most effective stages of disease. This evidence is critical for tailoring trials to specific AES subgroups and ensuring that future trials are adequately powered and targeted.

**Table 4:**
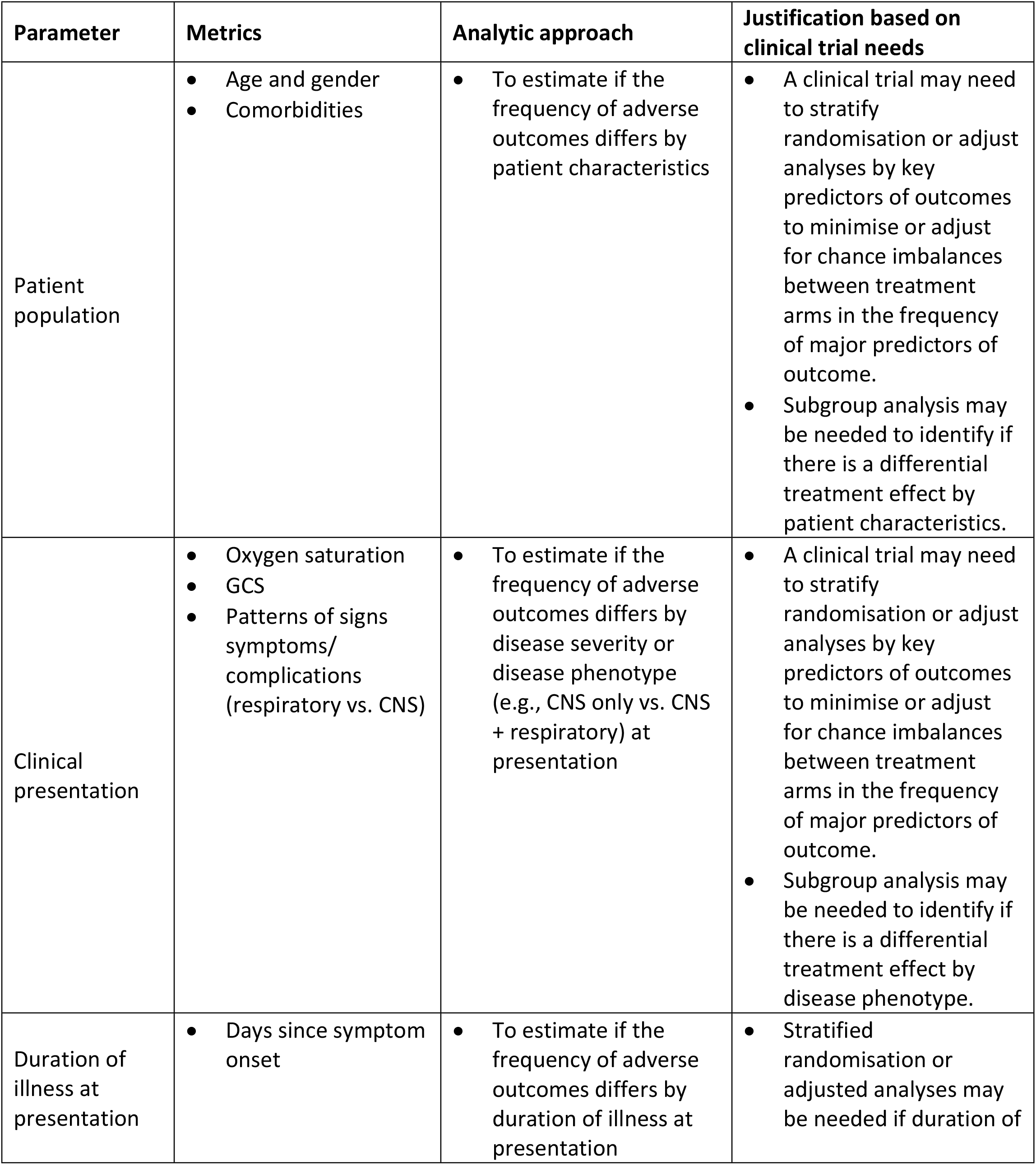
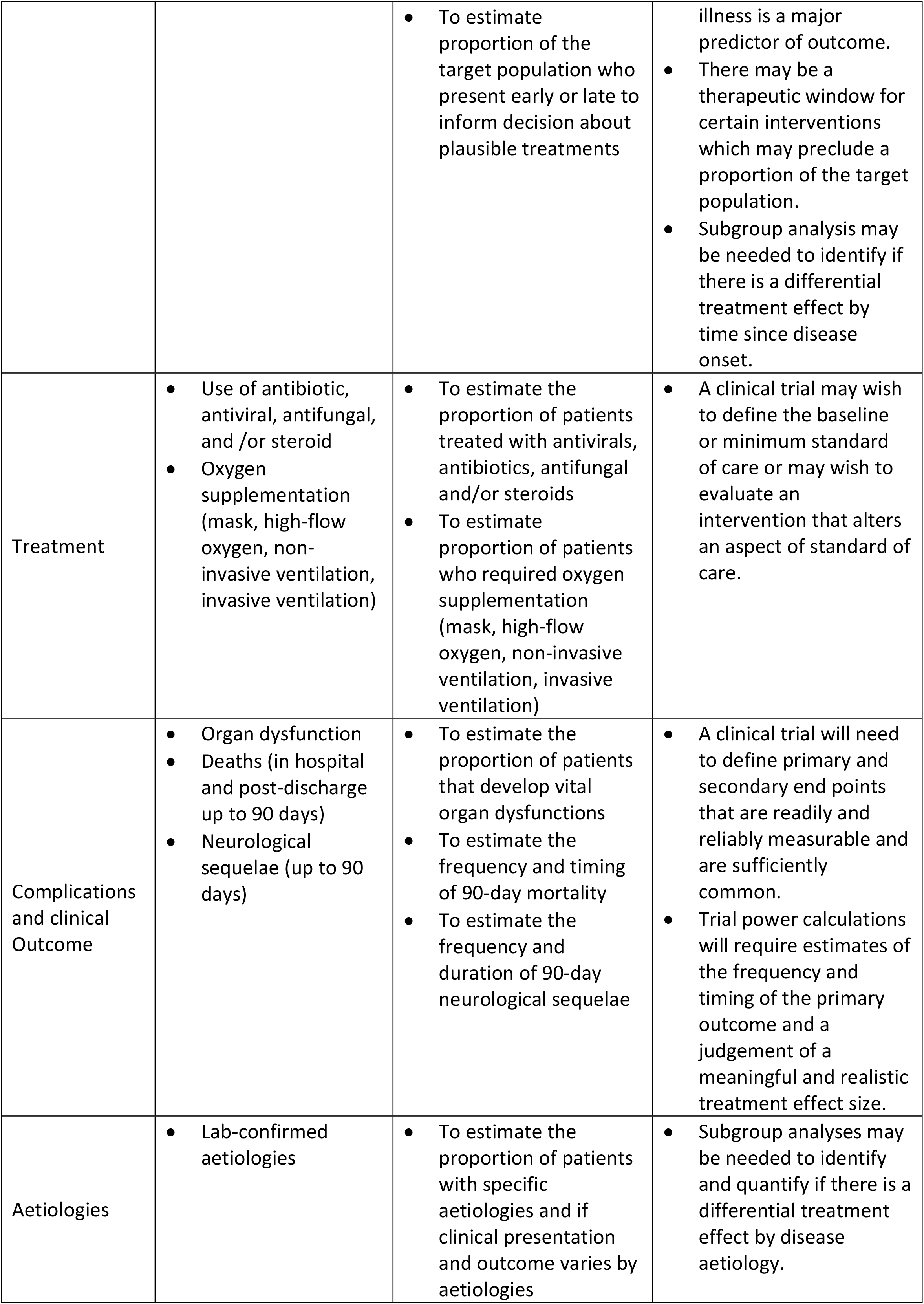
Key patient parameters, analysis framework and rationale based on clinical trial needs.

| Parameter | Metrics | Analytic approach | Justification based on clinical trial needs |
| --- | --- | --- | --- |
| Patient population | Age and gender<br>Comorbidities | To estimate if the frequency of adverse outcomes differs by patient characteristics | A clinical trial may need to stratify randomisation or adjust analyses by key predictors of outcomes to minimise or adjust for chance imbalances between treatment arms in the frequency of major predictors of outcome.<br>Subgroup analysis may be needed to identify if there is a differential treatment effect by patient characteristics. |
| Clinical presentation | Oxygen saturation<br>GCS<br>Patterns of signs symptoms/ complications (respiratory vs. CNS) | To estimate if the frequency of adverse outcomes differs by disease severity or disease phenotype (e.g., CNS only vs. CNS + respiratory) at presentation | A clinical trial may need to stratify randomisation or adjust analyses by key predictors of outcomes to minimise or adjust for chance imbalances between treatment arms in the frequency of major predictors of outcome.<br>Subgroup analysis may be needed to identify if there is a differential treatment effect by disease phenotype. |
| Duration of illness at presentation | Days since symptom onset | To estimate if the frequency of adverse outcomes differs by duration of illness at presentation | Stratified randomisation or adjusted analyses may be needed if duration of |
|  |  | To estimate proportion of the target population who present early or late to inform decision about plausible treatments | illness is a major predictor of outcome.<br>There may be a therapeutic window for certain interventions which may preclude a proportion of the target population.<br>Subgroup analysis may be needed to identify if there is a differential treatment effect by time since disease onset. |
| Treatment | Use of antibiotic, antiviral, antifungal, and /or steroid<br>Oxygen supplementation (mask, high-flow oxygen, non-invasive ventilation, invasive ventilation) | To estimate the proportion of patients treated with antivirals, antibiotics, antifungal and/or steroids<br>To estimate proportion of patients who required oxygen supplementation (mask, high-flow oxygen, non-invasive ventilation, invasive ventilation) | A clinical trial may wish to define the baseline or minimum standard of care or may wish to evaluate an intervention that alters an aspect of standard of care. |
| Complications and clinical Outcome | Organ dysfunction<br>Deaths (in hospital and post-discharge up to 90 days)<br>Neurological sequelae (up to 90 days) | To estimate the proportion of patients that develop vital organ dysfunctions<br>To estimate the frequency and timing of 90-day mortality<br>To estimate the frequency and duration of 90-day neurological sequelae | A clinical trial will need to define primary and secondary end points that are readily and reliably measurable and are sufficiently common.<br>Trial power calculations will require estimates of the frequency and timing of the primary outcome and a judgement of a meaningful and realistic treatment effect size. |
| Aetiologies | Lab-confirmed aetiologies | To estimate the proportion of patients with specific aetiologies and if clinical presentation and outcome varies by aetiologies | Subgroup analyses may be needed to identify and quantify if there is a differential treatment effect by disease aetiology. |

Data on treatment practices, including the use of antibiotics, antivirals, corticosteroids and oxygen supplementation, will provide insights into the current standard of care for AES. This will help establish baseline practices against which new interventions can be evaluated. The study will also document outcomes such as organ dysfunction, in-hospital mortality, and long-term neurological sequelae. These outcomes will inform the selection of primary and secondary endpoints for future trials, ensuring they are measurable, relevant, and common enough to provide meaningful results. Understanding the timing and frequency of these outcomes will be crucial for determining sample size and conducting power calculations for future trials.

While identifying the exact aetiology of AES is not the primary aim of the study, the collection of laboratory-confirmed diagnoses will help to identify common aetiologies. This will be useful in determining whether clinical presentations and outcomes vary by pathogen, which is crucial for future trials. Such data will enable subgroup analyses, assessing whether treatment effects differ based on the underlying cause of AES and aiding in managing the heterogeneity of the syndrome.

Several limitations may influence the study’s objectives. Missing data and unmeasured confounders, common in observational studies, could introduce bias and affect outcome estimates. Additionally, variability in diagnostic and care practices across hospitals may impact data consistency. However, the study incorporates rigorous data monitoring and multiple imputation techniques to mitigate these issues, ensuring the data’s robustness for future trial design.

The BASE study offers a valuable opportunity to overcome the limitations of conducting NiVD-specific trials by focusing on AES as a broader syndrome. Expanding this syndromic approach beyond Bangladesh to other regions with a high burden of infectious encephalitis could form the basis of a global network for harmonized data collection, similar to the International Severe Acute Respiratory and Emerging Infection Consortium (ISARIC)-WHO model for COVID-19 (37). Such a network would generate standardized, actionable data, optimizing clinical trial designs and improving patient outcomes for AES, including those affected by NiVD.

## Data Availability

No datasets were generated or analysed during the current study. All relevant data from this study will be made available upon study completion.

## List of Abbreviations

AES: Acute Encephalitis Syndrome
BASE: cohort Bangladesh Acute Encephalitis Syndrome cohort
CMV: Cytomegalovirus
CRF: Case Report Form
CSF: Cerebrospinal fluid
HSV: Herpes Simplex Virus
ICF: Informed Consent Form
IEDCR: Institute of Epidemiology, Disease Control, and Research
JE: Japanese Encephalitis
LFT: Liver function tests
NiV: Nipah virus
NiVD: Nipah virus disease
REC: Research Ethics Committee
RFT: Renal function test
SOP: Standard Operating Procedure

## Acknowledgment

MZH is a Moh Family Foundation Fellow at the Pandemic Sciences Institute, University of Oxford, and is also supported by scholarships from the Nuffield Department of Medicine, the Clarendon Fund, and the Reuben Foundation. PH holds the Moh Family Foundation Professorship of Emerging Infections and Global Health at the Pandemic Sciences Institute, University of Oxford. AR, LM, EG, JD, PH, and PO are supported by UK Inernational Development [301542-403], Wellcome [303666/Z/23/Z], and the Bill & Melinda Gates Foundation [INV-063472]. JK, CFS, TS, and JMM are supported by the US Centers for Disease Control and Prevention, Atlanta, USA. SS, MR, MHT, SY, FA, AFMP, MMR, ARMS, and TS are supported by the Ministry of Health and Family Welfare, Government of the People’s Republic of Bangladesh. DIR, MKMH, MEH, MWR, SSC, KIA, MK, MSIK, SB, SMS, and MZR received support from the icddr,b in Dhaka, Bangladesh; the organization is grateful to the Governments of Bangladesh and Canada for providing core unrestricted support. For the purpose of Open Access, the author has applied a CC BY public copyright license to any Author Accepted Manuscript version arising from this submission.

## Authors contributions

Md Zakiul Hassan (MZH) wrote the initial study protocol and drafted the manuscript. MZH and Amanda Rojek (AR) drafted the electronic Case Record Form (eCRF) and standard operating procedures (SOPs). AR, DIR, SS, MR, MKMU, MEH, MWR, LM, EG, JD, JB, SSC, KZ, MSIK, MHT, SY, FA, AFMP, MHR, ARMS, JDK, CFS, SB, JM, MZR, TS, SMS, PH, and PO contributed to the study design, critically reviewed the manuscript, and provided substantive feedback. All authors read and approved the final manuscript.

## Funding

The study is funded by Moh Family Foundation.

## Availability of data and materials

Recruitment is ongoing. The datasets will be made available upon the completion of recruitment and can be requested and shared in accordance with the data sharing policies of icddr,b and the International Severe Acute Respiratory and emerging Infection Consortium (ISARIC).

## Declarations

### Ethic’s approval and consent to participate

The Oxford Tropical Research Ethics Committee (OxTREC Ref: 576-23, approval date: January 25, 2024) and the Institutional Review Board (Research Review and Ethics Review Committee) of icddr,b (icddr,b Protocol Number: 24016, approval date: February 15, 2024) have reviewed and approved the study protocol, informed consent forms, and patient-facing materials for the BASE study.

### Consent for publication

Not applicable.

### Competing interests

The authors declare no competing interests.

### CDC disclaimer

The findings and conclusions in this report are those of the authors and do not necessarily represent the official position of the Centers for Disease Control and Prevention.

## Notes

### Competing Interest Statement

The authors have declared no competing interest.

### Funding Statement

Yes

### Author Declarations

The BASE study will be conducted in strict adherence to ethical standards, ensuring the confidentiality of all participants in accordance with applicable Good Clinical Practice (GCP) guidelines, national regulations, and the Declaration of Helsinki. Informed consent will be obtained from all participants or their legal representatives prior to enrolment. The consent process will clearly explain the study's purpose, procedures, potential risks and benefits, and the right to withdraw at any time without affecting future care. The consent form will be provided in Bengali, and participants will have ample opportunity to ask questions before and after deciding on participation. For those unable to read or write, the consent process will include an impartial witness to confirm comprehension and voluntary participation. To protect participant confidentiality, data will be pseudonymized, with each patient assigned a unique patient identification number, which will be consistently used throughout the study. This number will be assigned and managed securely by the centre coordinator, and personal data will only be released in accordance with local regulations and ethics committee approvals. The study received ethical approval from the Oxford Tropical Research Ethics Committee (OxTREC Ref: 576-23) and the Institutional Review Board (Research Review and Ethics Review Committee) of icddr,b (icddr,b Protocol Number: 24016). Any future amendments to the study protocol will also be submitted for review and approval to these committees.

